# Genome-Wide Association Study Identifies Novel Genetic Variants Associated with Widespread Pain in the UK Biobank (N=172,230)

**DOI:** 10.1101/2024.05.16.24307395

**Authors:** Qi Pan, Tengda Cai, Yiwen Tao, Roger Compte, Maryam Kazemi Naeini, Mainul Haque, Tania Dottorini, Frances M.K. Williams, Weihua Meng

## Abstract

**Objectives:** Widespread pain is a hallmark characteristic of fibromyalgia, commonly affecting older individuals. This study aimed to identify novel genetic variants associated with widespread pain by utilizing the extensive UK Biobank dataset.

**Methods:** We conducted a primary genome-wide association study (GWAS) using a novel definition of widespread pain, defined as pain experienced all over the body during the past month. Sex-stratified GWAS analysis approach was also performed to analyze the impact of sex on widespread pain.

**Results:** The primary GWAS identified one novel significant genetic locus (rs34691025, *p* = 1.76 × 10^-8^) on chromosome 5q13.2 within the *ARHGEF28* gene and several loci that approached genome-wide significance. The sex-stratified GWAS outputs revealed biological difference widespread pain between males and females, with a novel locus identified in the female-specific analysis within the *LRMDA* gene on chromosome 10. Genetic Correlation analysis demonstrated significant genetic correlations between widespread pain and other phenotypes, including joint disorders and spondylosis. The PheWAS revealed associations between the significant genetic variants with hearing disorders and cardiovascular diseases.

**Conclusions:** Our study advances the understanding of the genetic factors contributing to widespread pain, highlighting notable differences between males and females and identifying a novel genetic locus associated with this condition.

## Introduction

Widespread pain, commonly characterized by pain experienced in various parts of the body simultaneously, stands as a notable concern in clinical practice [1]. Unlike localized pain, which is confined to a specific region, widespread pain manifests across multiple regions, often without an apparent localized source. This type of pain is frequently observed in conditions such as fibromyalgia, rheumatoid arthritis, and certain neurological disorders, presenting a challenge in both diagnosis and treatment [2].

A commonly accepted definition of widespread pain is chronic widespread pain (CWP), which is characterized by the presence of widespread pain persisting for at least three months. A substantial body of epidemiological research suggests that the incidence and prevalence of CWP are significant, although these figures vary across different studies and countries. A systematic review indicates that the prevalence of CWP in the general population is approximately 10.6%, while in the UK, it is estimated to be 14.2% [3, 4]. The likelihood and risk factors of CWP increase with age, and females are more susceptible than males [4]. Beyond age and gender, CWP is associated with various risk factors, including sleep disorders, headaches and other types of pain, depression, and illness behavior [5]. The escalating issue of population aging has exacerbated the financial burden of CWP, leading to significant social costs. In the US, the direct and indirect annual expenses associated with CWP per patient are estimated to be approximately $12,428 [6]. Although the definition of CWP differs slightly from our focus on widespread pain, it is still meaningful to discuss the current epidemiological and genetic studies of CWP, as this information could reflect the nature of widespread pain.

The genetic basis of CWP has been the subject of extensive research, focusing particularly on its genetic architecture. Twin studies have played a pivotal role in dissecting the genetic and environmental contributions to CWP, with estimates indicating that genetic factors account for approximately 48–52% of the variance in CWP occurrence, underscoring a significant genetic component [7]. In a comprehensive GWAS meta-analysis that amalgamated data from 14 studies, a notable locus was identified on chromosome 5, positioned intergenically between *CCT5* and *FAM173B* [8]. Furthermore, a recent study by Rahman et al. utilizing the UK Biobank data on CWP identified the *RNF123* locus as being associated with CWP [9]. This association was specifically observed in cases reporting pain all over the body and persisting for over three months.

Given the extensive research on CWP, our study shifts the focus to widespread pain experienced within the last month, a criterion less explored in the literature. This unique definition allows for a more precise investigation of the genetic underpinnings of short-term widespread pain, which may differ from those of chronic conditions. The aim of this study is to identify novel genetic variants linked to widespread pain in the UK Biobank by conducting the GWAS research. We employ a unique definition that considers pain experienced all over the body during the past month for the GWAS and utilize three independent replication datasets (FibroGene, FinnGen and TwinsUK). Sex-stratified GWAS analysis approach is firstly conducted to analyze the impact of sex on widespread pain. Through this research, we hope to identify new loci associated with widespread pain, thereby uncovering novel genetic mechanisms and offering fresh insights for the treatment of this condition.

## Methods

### Cohorts’ information

The UK Biobank was employed as the discovery cohort in our study. This cohort consists of over 500,000 participants aged 40-69 from England, Scotland, and Wales, recruited between 2006 and 2011. They underwent clinical examinations, completed comprehensive questionnaires, and provided DNA samples with informed consent for research use. The study received ethical approval from the UK’s National Health Service National Research Ethics Service (reference 11/NW/0382). Further details can be found at www.ukbiobank.ac.uk.

The DNA extraction and quality control (QC) processes were standardized, with detailed methodologies available at https://biobank.ctsu.ox.ac.uk/crystal/ukb/docs/genotyping_sample_workflow.pdf. The Welcome Trust Centre for Human Genetics at Oxford University oversaw the standardized QC for genotyping results, with a comprehensive description at http://biobank.ctsu.ox.ac.uk/crystal/refer.cgi?id=155580. In July 2017, the UK Biobank released genetic data, including genotyped and imputed genotypes, from 501,708 samples to authorized researchers. Bycroft et al. provided a detailed description of the QC procedures for imputation [10].

In this study, a specific pain-related question developed by the UK Biobank was utilized: “In the last month, have you experienced any of the following that interfered with your usual activities?” Participants could select from the following options: (1) Headache; (2) Facial pain; (3) Neck or shoulder pain; (4) Back pain; (5) Stomach or abdominal pain; (6) Hip pain; (7) Knee pain; (8) Pain all over the body; (9) None of the above; (10) Prefer not to say (UK Biobank Questionnaire field ID: 6159). Multiple selections were allowed. Cases of widespread pain were identified by the selection of “Pain all over the body,” regardless of other choices. Controls were those who chose “None of the above.” The analysis excluded data from individuals not of white British descent (UK Biobank Questionnaire field ID: 21000) and those with self-reported diagnoses of rheumatoid arthritis, polymyalgia rheumatica, unspecified arthritis, systemic lupus erythematosus, ankylosing spondylitis, and myopathy (UK Biobank Questionnaire field ID: 20002).

In the replication phase, we employed publicly available summary statistics for the phenotype of fibromyalgia as classified by ICD-10 in the FinnGen dataset, as well as summary statistics from the FibroGene fibromyalgia study and the CWP study within TwinsUK. The FinnGen dataset comprised 3,761 cases diagnosed with fibromyalgia and 375,928 controls without the diagnosis. The GWAS for the FinnGen cohort was conducted according to the protocols of FinnGen GWAS round 9, with details available at https://finngen.gitbook.io/documentation [11]. Detailed information on sample phenotyping, genotyping, and the GWAS methodology applied to the FinnGen sample can be found at https://risteys.finngen.fi/ [11]. The FibroGene dataset included 905 fibromyalgia cases recruited via advertisements in Fibromyalgia Action UK charity resources, and 14,213 controls from the general population program of the UK BioResource project. Details on genotyping and GWAS methodology for the FibroGene dataset can be referenced at https://bioresource.nihr.ac.uk/media/nlybpdcd/uk_axiom_biobank_genotyping_arrays_datasheet.pdf [12]. The TwinsUK GWAS study focusing on CWP comprised 903 individuals with CWP and 2,369 control subjects The TwinsUK cohort, established as a registry of volunteer same-sex twins across the United Kingdom, encompasses a broad age range from 16 to 98 years. Initiated in 1992, the cohort has been continuously gathering a wealth of data and biological materials from its 14,274 registered twins. The ethical integrity of the study is upheld through approval from the Research Ethics Committee at Guy’s and St. Thomas’ NHS Foundation Trust. A significant aspect of the TwinsUK cohort is its repository of genetic information from 6,921 participants. This genetic data has undergone quality control, documented by Moayyeri et al [13]. For SNPs missing in the replication datasets, we employed a strategy based on linkage disequilibrium and physical distance to identify and use the most appropriate SNP for replication. Additionally, it is important to note that the replication *p*-values have not been adjusted for multiple testing across the 14 genes tested for replication.

### GWAS and Statistical analysis

Our study comprised three independent GWAS. The primary GWAS was conducted with the objective of understanding the genetic foundation of widespread pain across the entire dataset. The secondary GWAS was designed to assess the influence of gender on widespread pain, with the analysis specifically focused on conducting separate GWAS for each sex. In this study, the Genome-wide Complex Trait Analysis (GCTA, v1.94.1) software (available at https://yanglab.westlake.edu.cn/software/gcta/#Overview)) was employed as the principal tool for executing GWAS [14]. Our research utilized the fastGWA function within GCTA, a mixed linear model association tool, for the GWAS analyses. Standard QC procedures were implemented, which involved the exclusion of single nucleotide polymorphisms (SNPs) with INFO scores below 0.3, SNPs with minor allele frequencies lower than 0.5%, or SNPs failing the Hardy-Weinberg tests (*p* < 10^-6^). Furthermore, SNPs located on the X and Y chromosomes, as well as mitochondrial SNPs, were excluded from the analysis. The association tests were conducted using the fastGWA function, with adjustments for age, sex, body mass index (BMI), and eight population principal components. A *χ*^2^ test was utilized to assess gender differences between the cases and controls, while the comparison of age and BMI was performed using an independent t-test, employing R v4.2.2. R v4.2.2 was also used to select the data of white British descent in UK Biobank data and divide the cases and controls of widespread pain. A *p-value* less than 5 x 10^-8^ was considered indicative of genome-wide association significance. Additionally, GCTA was also employed to compute the narrow-sense heritability.

### GWAS-associated analysis by FUMA and LDSC

The FUMA web application served as the primary annotation tool throughout widespread pain study [15]. Additionally, we generated both Manhattan and Q-Q plots using this application. For regional visualization, we employed LocusZoom (http://locuszoom.org/) [16]. Notably, FUMA facilitated three key types of analyses, gene analysis, gene-set analysis, and tissue expression analysis. In the gene analysis, the summary statistics of SNPs were aggregated to the level of entire genes, allowing for the assessment of associations between genes and the phenotype under investigation. In the gene-set analysis, specific groups of genes sharing common biological, functional, or other characteristics were collectively tested, providing insights into the involvement of distinct biological pathways or cellular functions in the genetic basis of the observed phenotype. The tissue expression analysis was conducted using data from GTEx (https://www.gtexportal.org/home), which has been integrated into the FUMA platform. For this analysis, the average gene expression per tissue type was used as a gene covariate, enabling the examination of relationships between gene expression in specific tissue types and the genetic associations with widespread pain, the focus of our study. We generated two gene expression heatmaps for genes identified through positional mapping. These heatmaps represent the average of normalized expression values, employing data from GTEx version 8 across 54 and 30 general tissue types. Tissue specificity was assessed by examining the differentially expressed genes (DEG) for each tissue type. Furthermore, an enrichment test for DEGs was performed for both sets of tissue types in GTEx version 8, focusing specifically on genes identified through positional mapping rather than utilizing the full distribution of SNP p-values in MAGMA tissue expression analysis.

To explore potential genetic correlations between widespread pain and various other phenotypes, we assessed the genetic associations of widespread pain with 1,396 characteristics from the UK Biobank through the Complex Trait Virtual Lab (CTG-VL) (https://genoma.io/). CTG-VL is an open-source platform that integrates available GWAS datasets to enable the estimation of genetic correlations for complex traits. Subsequently, to further investigate any potential genetic differences between the sexes, we utilized linkage disequilibrium score regression (LDSC) via LDSCv1.0.1 (available at https://github.com/bulik/ldsc) to examine the genetic correlation of widespread pain between males and females. LDSC utilizes the principle of linkage disequilibrium, which refers to the non-random association of alleles at different loci within a population [17].

### Expression quantitative trait loci (eQTL), chromatin interaction analysis and positional mapping

Expression quantitative trait loci (eQTL) have become instrumental in elucidating the regulatory mechanisms of variants identified through GWAS. Cis-eQTL notably impact gene expression by interacting with variants that are situated in proximity (within 1 Mb) to the gene [18]. In eukaryotic cells, the genome is compactly organized within the micron-sized nucleus, with chromatin serving as the fundamental structural unit. This organization is essential, arranging the genome into a three-dimensional structure that is pivotal for processes such as DNA replication, DNA damage repair, gene transcription, and other crucial biological functions [19]. Positional mapping within our study was conducted using a maximum distance of 10 kb. The integration of cis-eQTL analysis, chromatin interaction analysis, and positional mapping in our gene mapping approach offers a comprehensive perspective on the genomic architecture underpinning these findings.

### Phenome-wide association analysis (PheWAS)

Our study implemented a Phenome-Wide Association Analysis (PheWAS) to investigate the relationships between significant SNP associations, their corresponding genes, and multiple phenotypes. The primary objectives of this analysis were to corroborate the associations identified in our GWAS with pain phenotypes and to uncover new associations between genetic variants linked to widespread pain and other phenotypes. The PheWAS was conducted utilizing an expansive dataset comprising 4,756 GWAS summary statistics available on the GWAS ATLAS platform https://atlas.ctglab.nl/PheWAS [20]. For this analysis, only SNPs exhibiting associations with *p*-values lower than 0.05 were included. Furthermore, adjustments for multiple comparisons were performed using the Bonferroni correction method.

## Results

### GWAS results

During the initial assessment phase (2006-2010) of the UK Biobank study, a total of 501,708 participants were administered a pain questionnaire. In our research, we excluded non-white British participants and samples that failed to meet QC criteria. In the primary GWAS, we identified 4,617 cases (comprising 1,885 males and 2,732 females) and 167,613 controls (consisting of 80,228 males and 87,385 females) for analysis. The study utilized 11,165,459 SNPs for the GWAS examination. The secondary phase of the study involved sex-stratified GWAS analyses, where, following identical QC procedures, the female cohort (90,117 samples) consisted of 2,732 cases and 87,385 controls, while the male cohort (82,113 samples) included 1,885 cases and 80,228 controls. Table 1 provides a comprehensive summary of the clinical characteristics of cases and controls in the primary GWAS. Table 2 presents a detailed overview of the cases and controls in the secondary sex-stratified GWAS.

**Table 1.** Clinical characteristics of widespread pain cases and controls in the UK Biobank.

| Covariates | Cases | Controls | *p*-value |
| --- | --- | --- | --- |
| Sex (male: female) | 1,885:2,732 | 80,228:87,385 | <0.001 |
| Age (years) | 56.7 (7.67) | 56.9 (7.95) | 0.55 |
| BMI (kg/m<sup>3</sup>) | 29.6 (5.77) | 26.7 (4.28) | <0.001 |

A $\chi^2$ test was used to test the difference of gender frequency between cases and controls and an independent t test was used for other covariates.
Continuous covariates were presented as mean (standard deviation)
BMI represents body mass index.

**Table 2.** Clinical characteristics of widespread pain cases and controls in the UK Biobank (sex-stratified GWAS)

| GWAS Analysis | Covariates | Cases | Controls | *p*-value |
| --- | --- | --- | --- | --- |
| Male-specific GWAS | Age (years) | 57.3 (7.60) | 57.2 (8.04) | 0.054 |
|  | BMI (kg/m<sup>3</sup>) | 29.6 (5.11) | 27.3 (3.90) | <0.001 |
| Female-specific GWAS | Age (years) | 57.2 (7.58) | 56.8 (7.87) | <0.05 |
|  | BMI (kg/m<sup>3</sup>) | 29.8 (6.36) | 26.2 (4.57) | <0.001 |

Total number of females: 90,117; Total number of males: 82,113. An independent t test was used for other covariates.
Continuous covariates were presented as mean (standard deviation)
BMI represents body mass index.

In our study, we identified one novel and distinct SNP cluster on chromosome 5 that exhibited significant associations with widespread pain, achieving genome-wide significance (*p* < 5 × 10^-8^), as delineated in Figure 1. Additionally, we identified several significant loci proximal to the genome-wide significant threshold, which are listed in Table 3. This table enumerates a total of 14 SNPs, encompassing both significant and marginally significant SNPs, with each representing the most statistically significant association within its respective locus. A detailed enumeration of all significantly associated SNPs from this GWAS is furnished in Supplementary Table 1. Notably, the significant association was observed in the SNP cluster situated within the *ARHGEF28* gene on chromosome 5q13.2, with a *p-*value of 1.76 × 10^-8^ for rs34691025. The regional plot illustrating the most significant locus in *ARHGEF28* is provided in Figure 2. Moreover, the Q-Q plot of the GWAS during the discovery phase is presented in Figure 3. The SNP-based heritability for widespread pain was estimated to be 0.18, with a standard error of 0.02.

**Fig.1.**
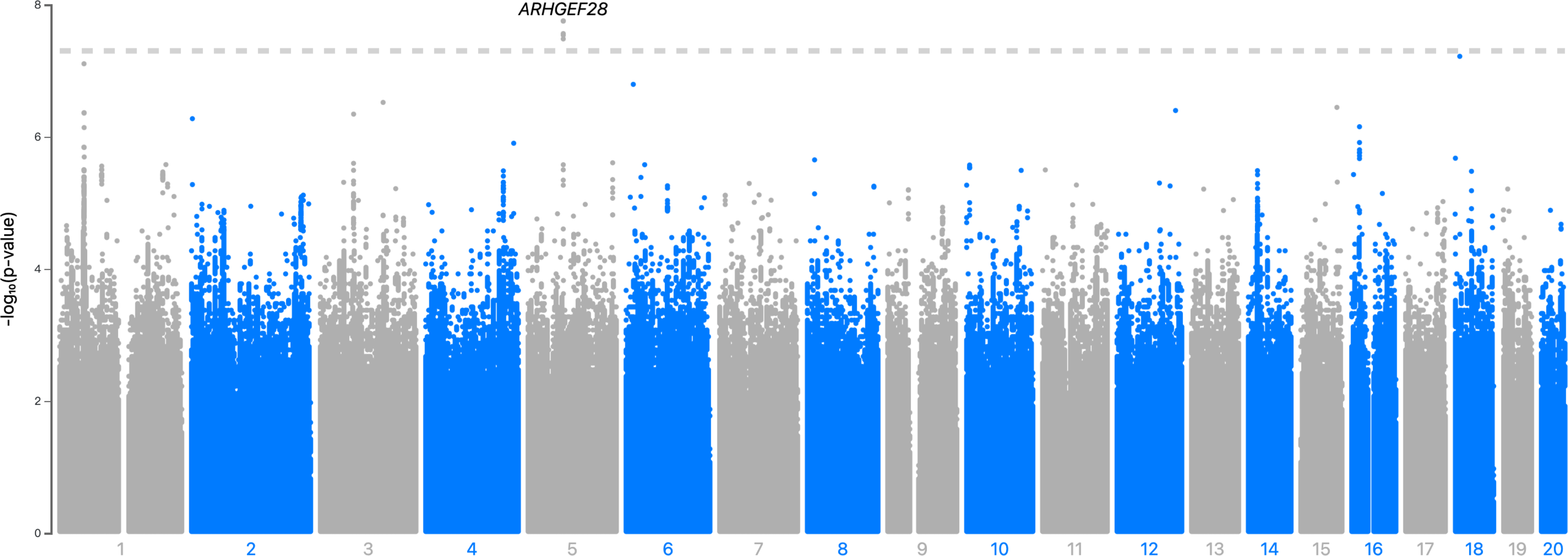
The Manhattan plot of the GWAS analysis on widespread pain (N = 172,230) The dashed grey line indicates the cut-off *p-*value of 5 × 10^−8^

**Fig.2.**
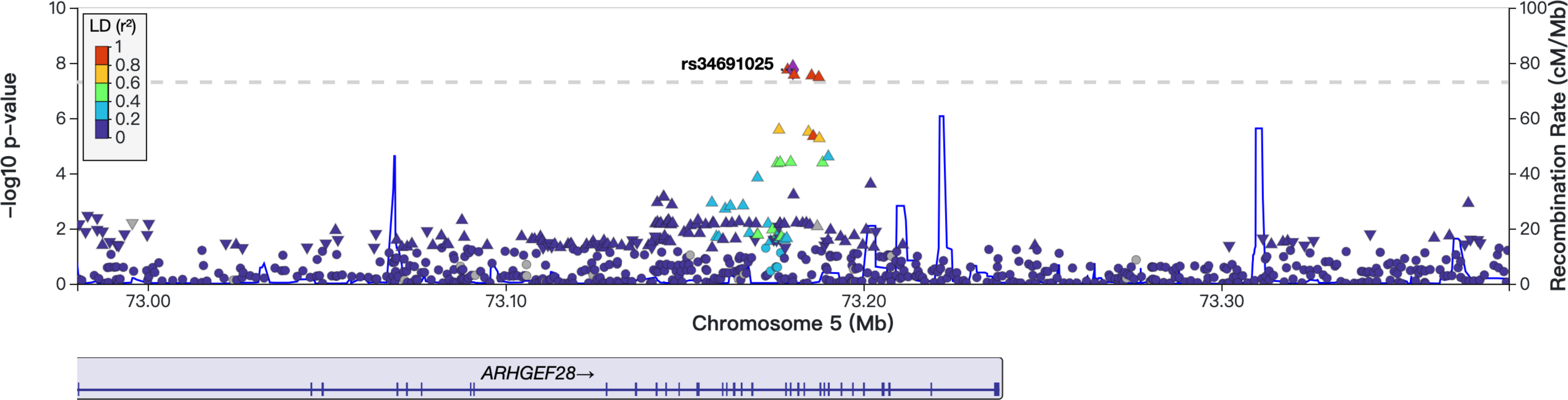
The regional plots of locus in *ARHGEF28* region

**Fig.3.**
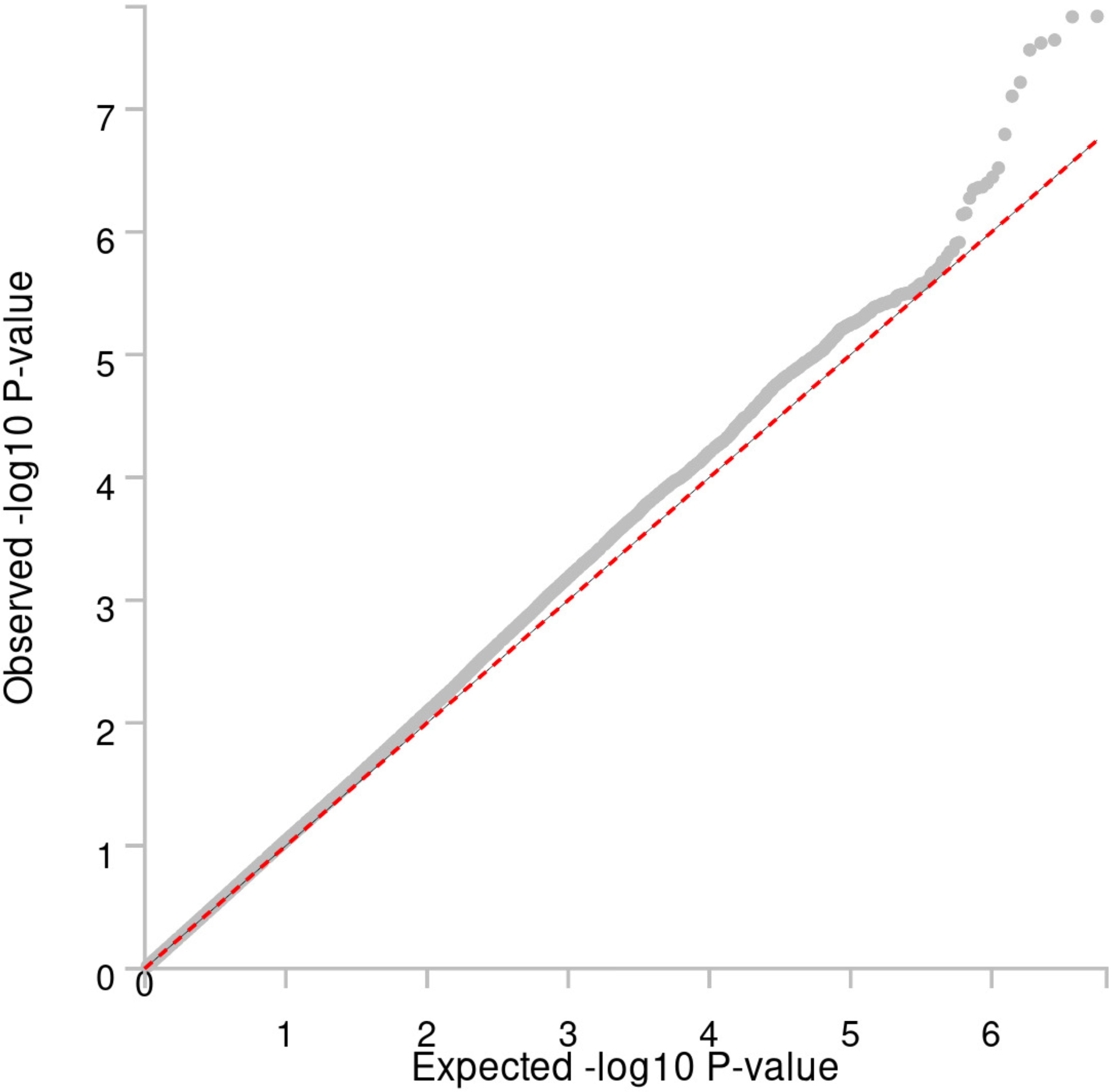
The Q-Q plot of the GWAS analysis on widespread pain (N = 172,230)

**Table 3.**
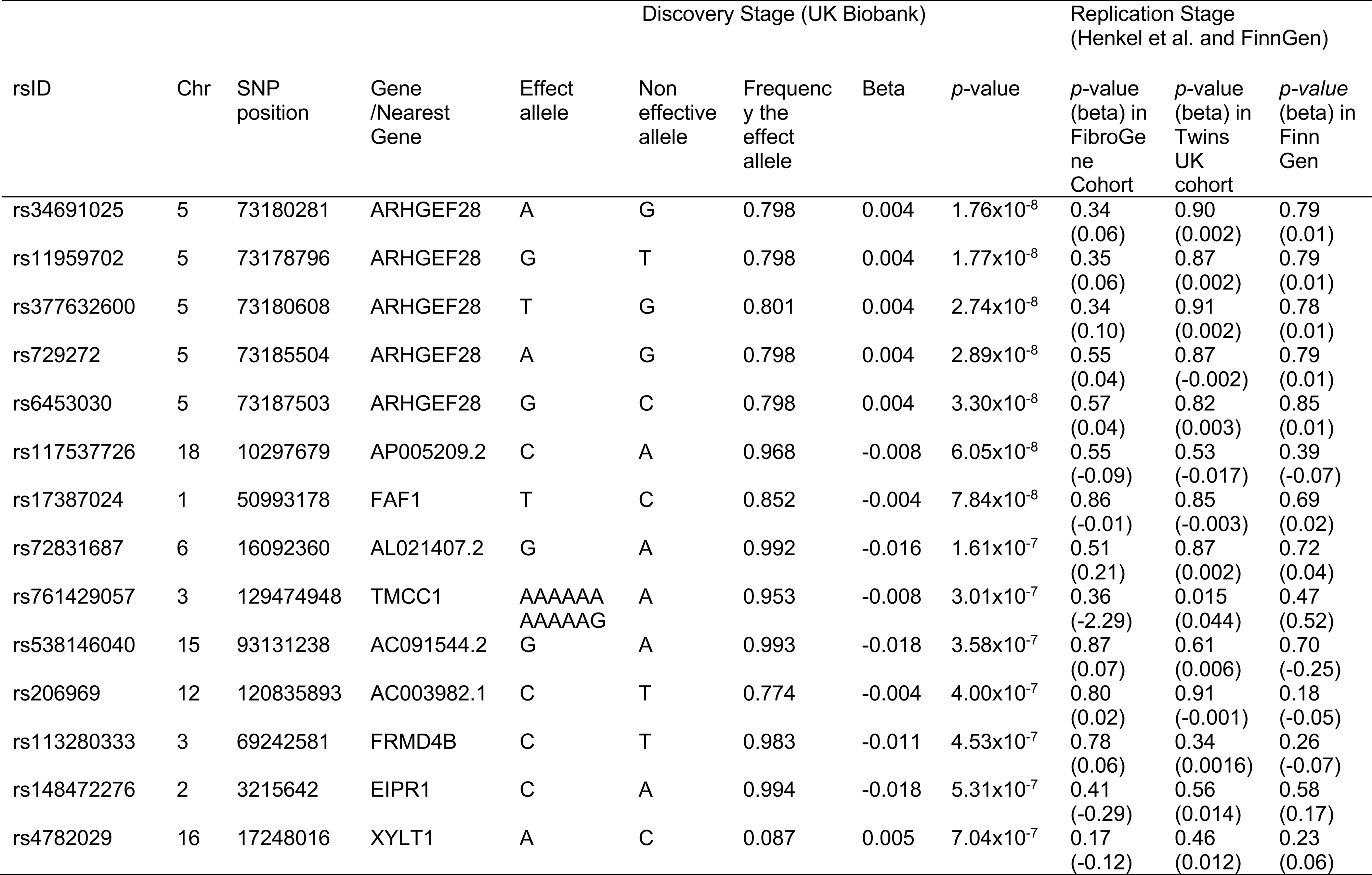
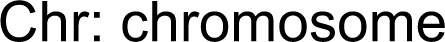
The Novel Significant SNP and SNPs Approaching Significance Identified by GWAS on Widespread Pain.

| rsID | Chr | SNP position | Gene /Nearest Gene | Effect allele | Discovery Stage (UK Biobank) |  |  |  | Replication Stage (Henkel et al. and FinnGen) |  |  |
| --- | --- | --- | --- | --- | --- | --- | --- | --- | --- | --- | --- |
|  |  |  |  |  | Non effective allele | Frequency the effect allele | Beta | p-value | p-value (beta) in FibroGene Cohort | p-value (beta) in Twins UK cohort | p-value (beta) in Finn Gen |
| rs34691025 | 5 | 73180281 | ARHGEF28 | A | G | 0.798 | 0.004 | 1.76x10<sup>-8</sup> | 0.34<br>(0.06) | 0.90<br>(0.002) | 0.79<br>(0.01) |
| rs11959702 | 5 | 73178796 | ARHGEF28 | G | T | 0.798 | 0.004 | 1.77x10<sup>-8</sup> | 0.35<br>(0.06) | 0.87<br>(0.002) | 0.79<br>(0.01) |
| rs377632600 | 5 | 73180608 | ARHGEF28 | T | G | 0.801 | 0.004 | 2.74x10<sup>-8</sup> | 0.34<br>(0.10) | 0.91<br>(0.002) | 0.78<br>(0.01) |
| rs729272 | 5 | 73185504 | ARHGEF28 | A | G | 0.798 | 0.004 | 2.89x10<sup>-8</sup> | 0.55<br>(0.04) | 0.87<br>(-0.002) | 0.79<br>(0.01) |
| rs6453030 | 5 | 73187503 | ARHGEF28 | G | C | 0.798 | 0.004 | 3.30x10<sup>-8</sup> | 0.57<br>(0.04) | 0.82<br>(0.003) | 0.85<br>(0.01) |
| rs117537726 | 18 | 10297679 | AP005209.2 | C | A | 0.968 | -0.008 | 6.05x10<sup>-8</sup> | 0.55<br>(-0.09) | 0.53<br>(-0.017) | 0.39<br>(-0.07) |
| rs17387024 | 1 | 50993178 | FAF1 | T | C | 0.852 | -0.004 | 7.84x10<sup>-8</sup> | 0.86<br>(-0.01) | 0.85<br>(-0.003) | 0.69<br>(0.02) |
| rs72831687 | 6 | 16092360 | AL021407.2 | G | A | 0.992 | -0.016 | 1.61x10<sup>-7</sup> | 0.51<br>(0.21) | 0.87<br>(0.002) | 0.72<br>(0.04) |
| rs761429057 | 3 | 129474948 | TMCC1 | AAAAAA<br>AAAAAG | A | 0.953 | -0.008 | 3.01x10<sup>-7</sup> | 0.36<br>(-2.29) | 0.015<br>(0.044) | 0.47<br>(0.52) |
| rs538146040 | 15 | 93131238 | AC091544.2 | G | A | 0.993 | -0.018 | 3.58x10<sup>-7</sup> | 0.87<br>(0.07) | 0.61<br>(0.006) | 0.70<br>(-0.25) |
| rs206969 | 12 | 120835893 | AC003982.1 | C | T | 0.774 | -0.004 | 4.00x10<sup>-7</sup> | 0.80<br>(0.02) | 0.91<br>(-0.001) | 0.18<br>(-0.05) |
| rs113280333 | 3 | 69242581 | FRMD4B | C | T | 0.983 | -0.011 | 4.53x10<sup>-7</sup> | 0.78<br>(0.06) | 0.34<br>(0.0016) | 0.26<br>(-0.07) |
| rs148472276 | 2 | 3215642 | EIPR1 | C | A | 0.994 | -0.018 | 5.31x10<sup>-7</sup> | 0.41<br>(-0.29) | 0.56<br>(0.014) | 0.58<br>(0.17) |
| rs4782029 | 16 | 17248016 | XYLT1 | A | C | 0.087 | 0.005 | 7.04x10<sup>-7</sup> | 0.17<br>(-0.12) | 0.46<br>(0.012) | 0.23<br>(0.06) |

Chr: chromosome

We tested the significant and independent SNPs identified in our discovery analysis, as well as other SNPs proximal to the genome-wide significance threshold, for replication in the FinnGen, FibroGene, and Twins UK cohorts. Within the TwinsUK cohort, specifically regarding CWP, we observed weak replication for rs761429057 in the *TMCC1* gene (*p* = 0.017) among the 14 SNPs from the discovery cohort. However, in the context of fibromyalgia within the FibroGene and FinnGen cohorts, none of the 14 SNPs demonstrated replication in either cohort (*p* > 0.05). The *p-*values of the associations of these 14 independent SNPs from the discovery stage were extracted from FinnGen, FibroGene and Twins UK cohorts are presented in Table 3.

In the sex-stratified GWAS analyses, the male-specific GWAS did not identify any single locus of genome-wide significance associated with widespread pain. Conversely, the female-specific GWAS revealed one significant locus associated with widespread pain, which differed from the findings of the primary GWAS. This novel locus was identified within the *LRMDA* gene on chromosome 10, with a *p* = 4.04 × 10^-8^ for rs137998089. Detailed information regarding these findings is provided in Table 4, and the corresponding Manhattan plot and regional plot are depicted in Figure 4 and 5, respectively.

**Fig.4.**
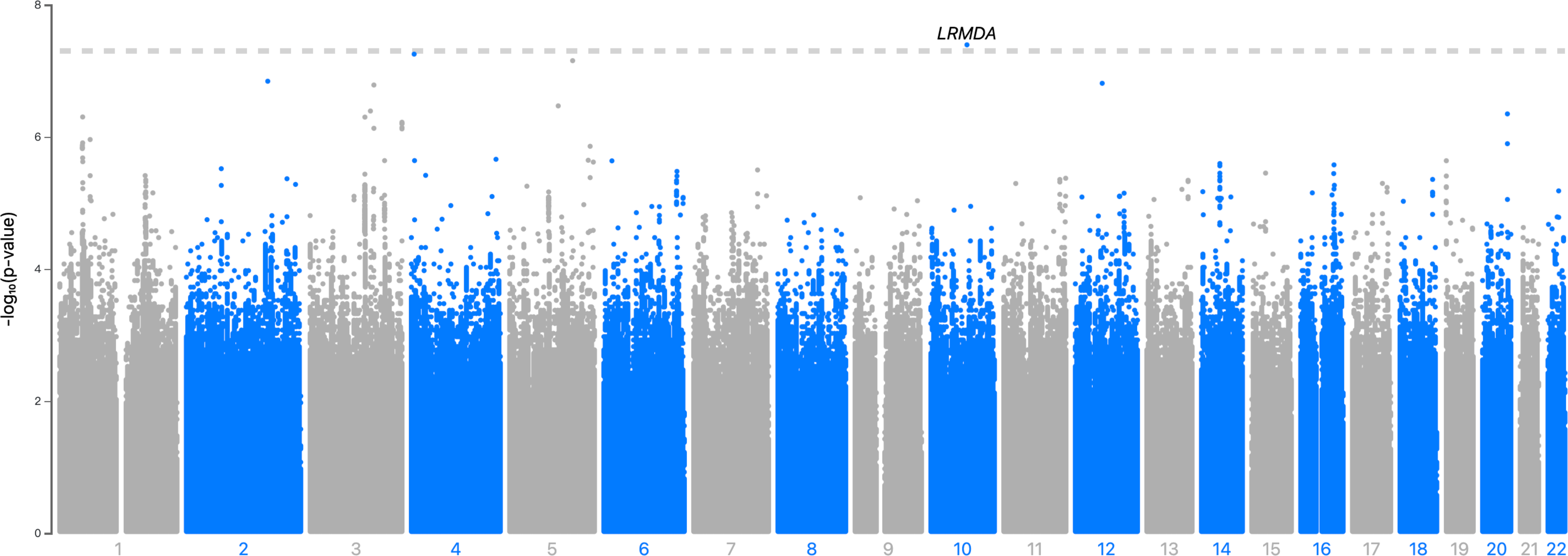
The Manhattan plot of the female-specific GWAS analysis on widespread pain (N = 90,117) The dashed grey line indicates the cut-off *p-*value of 5 × 10^−8^

**Fig.5.**
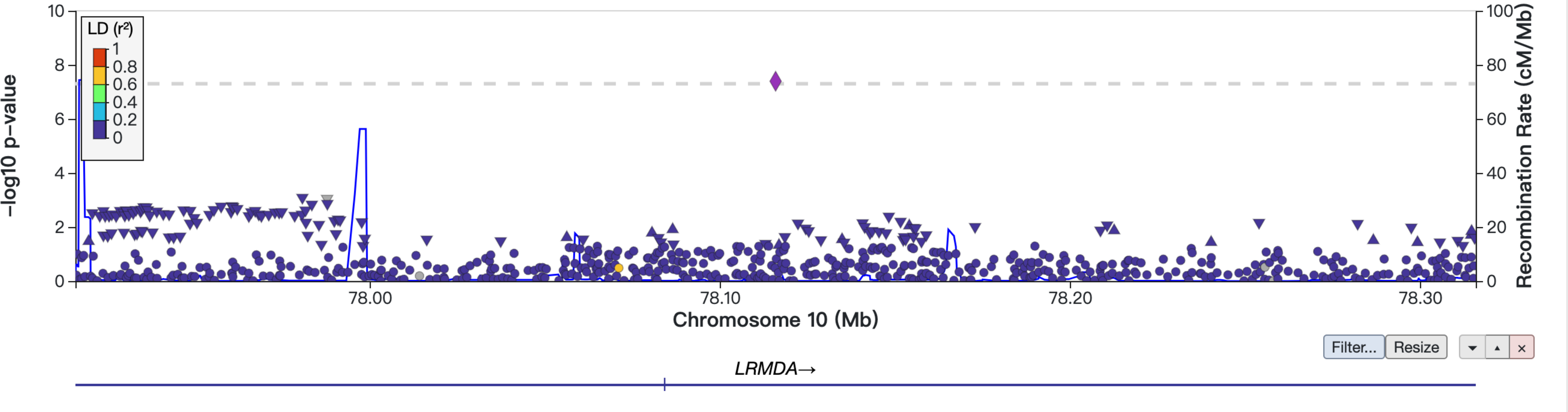
The regional plots of locus in *LRMDA* region

**Table 4.** The locus and top SNP identified by the female-specific GWAS on widespread pain.

| Locus Rank | rsID | Chr | SNP position | Nearest Gene | Effect allele | Non effective allele | Frequency the effect allele | *p* value | beta |
| --- | --- | --- | --- | --- | --- | --- | --- | --- | --- |
| 1 | rs137998089 | 10 | 78115898 | *LRMDA* | A | G | 0.994 | 4.03x10<sup>-8</sup> | -0.003 |

Chr: chromosome

### Gene, gene-set and tissue expression analysis

In our gene analysis, we mapped SNPs within genes to a total of 18,115 protein-coding genes. As a result, we established a genome-wide significance threshold of *p* < 0.05 / 18,115 = 2.76 × 10^-6^. At this stage, no genes met the criteria for significance. In our analysis of gene sets, we evaluated 17,109 gene sets in total. The threshold for statistical significance on a genome-wide scale was thus set at *p* < 0.05 / 17,109 = 2.92 × 10^-6^. However, none of the gene sets reached statistical significance. The top ten gene sets identified in this analysis are listed in Supplementary Table 2.

In the tissue expression analysis, we identified 30 general tissue types and 53 specific tissue types. However, none of the tissues exhibited a significant association. Further details and visualization of these results are available in Supplementary Figure 1 and 2.

In the gene expression heatmaps presented in Figure 6, the gene *ARHGEF28* exhibits higher expression levels in the nerve, oral, and adrenal gland tissues compared to other tissue types. Tissue specificity testing did not yield significant differences in the DEG across both the 54 and 30 tissue types analyzed in GTEx version 8. Detailed visualizations of these findings are included in Supplementary Figure 3.

**Fig.6.**
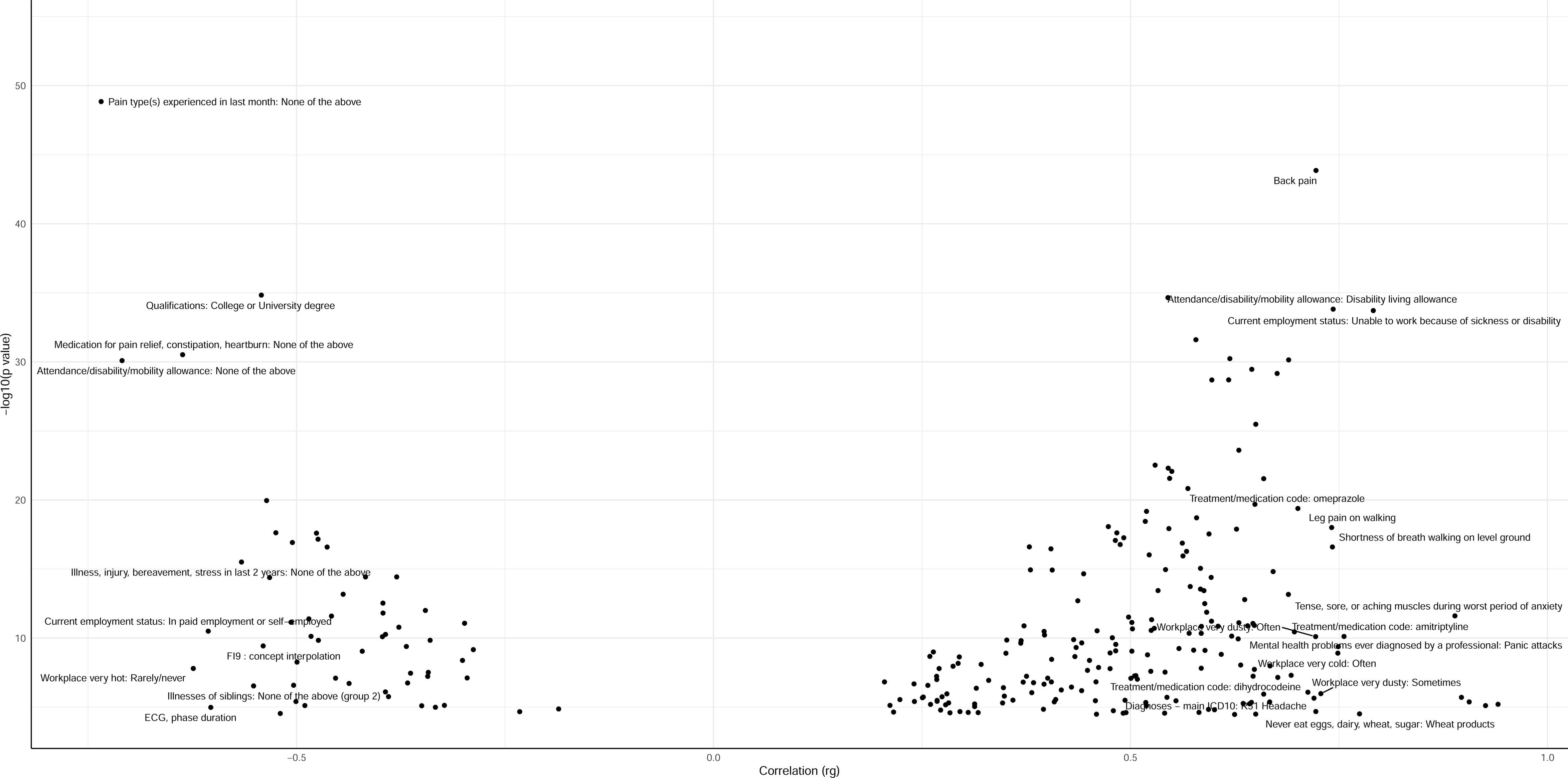
Gene expression heatmaps across 54 tissue types and 30 general tissue Categories from GTEx Version 8

### Genetic correlation analysis by LDSC

In our study examining the relationship between widespread pain and various pain phenotypes, as well as the genetic correlation between widespread pain in females and males, we discovered several significant genetic correlations, as depicted in Figure 7. After correcting for multiple testing, the three most notable genetic correlations (*r*_*g*_) were found in the following categories: other joint disorders, not elsewhere classified (*r*_*g*_ = 0.94, *p* = 6.20 × 10^-6^), spondylosis (*r*_*g*_ = 0.93, *p* = 7.73 × 10^-6^), and other diseases of the anus and rectum (*r*_*g*_ = 0.89, *p* = 1.95 × 10^-6^). The genetic correlation between widespread pain in females and males was determined to be *r*_*g*_= 0.80 (*p* = 7.36 × 10^-92^). The comprehensive results are provided in Supplementary Table 3.

**Fig.7.**
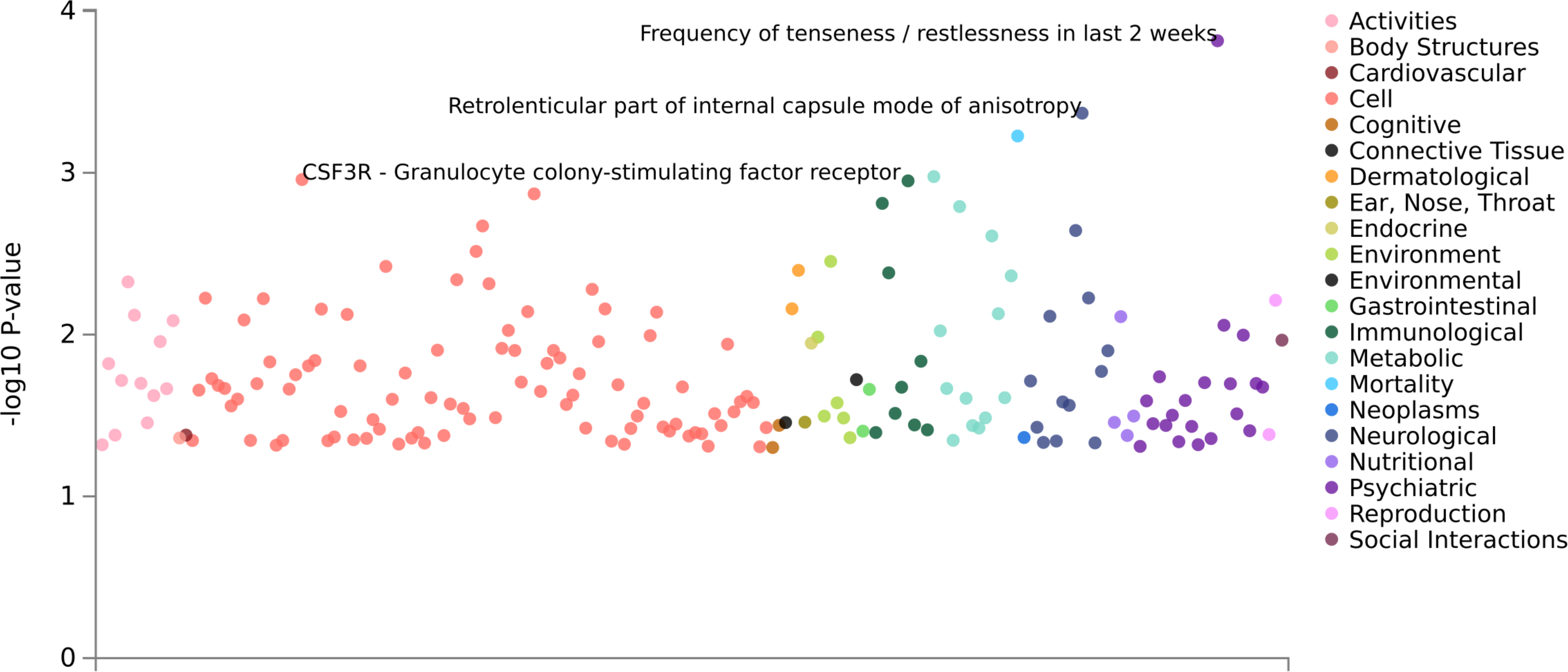
Genetic correlation results for widespread pain using LDSC on CTG-VL. For a full list, see Supplementary Table 3

### Expression quantitative trait loci (eQTL), chromatin interaction analysis and positional mapping

From the cis-eQTL analysis, SNPs rs35588745 and rs71636094 were found to be significantly associated with the tissue type Cells_Cultured_fibroblasts, achieving a False Discovery Rate (FDR) of less than 0.05. The results of the chromatin interactions and cis-eQTL analyses are illustrated in the circus plot presented in Figure 8. This plot shows the genes that interact with *ARHGEF28*, including *IQGAP2, GCTN4, FAM169A, NSA2, GFM2, HEXB, ENC1, UTP14*, and *AC008387.1*. Additionally, the positional mapping analysis did not identify significant results.

**Fig.8.**
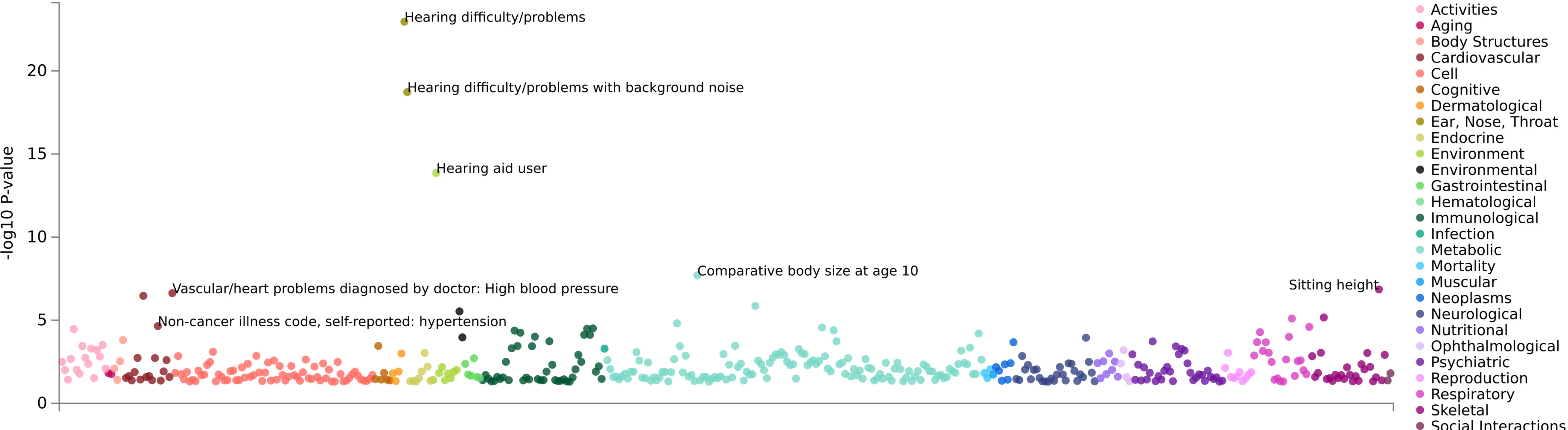
Circos plot illustrating chromatin interactions and eQTLs on chromosome 5

### PheWAS

PheWAS were conducted using the GWAS ATLAS platform to explore phenotypes associated with independently significant SNPs (rs34691025) and significantly associated genes (*ARHGEF28*). SNP rs34691025 exhibited significant associations with the frequency of tenseness/restlessness in the last two weeks (*p* = 1.5 × 10^-4^), the mode of anisotropy in the retrolenticular part of the internal capsule (*p* = 1.70 × 10^-6^), and long-standing illness, disability, or infirmity (*p* = 3.60 × 10^-12^). The gene *ARHGEF28* was associated with hearing difficulty/problems (*p* = 1.09 × 10^-23^), high blood pressure (*p* = 3.42 × 10^-7^), and hypertension (*p* = 2.26 × 10^-5^). All significant traits that passed the Bonferroni correction are listed in Supplementary Table 4. The plots illustrating phenotypes associated with the SNP and gene are visualized in Figure 9 and 10, respectively.

**Fig.9.**
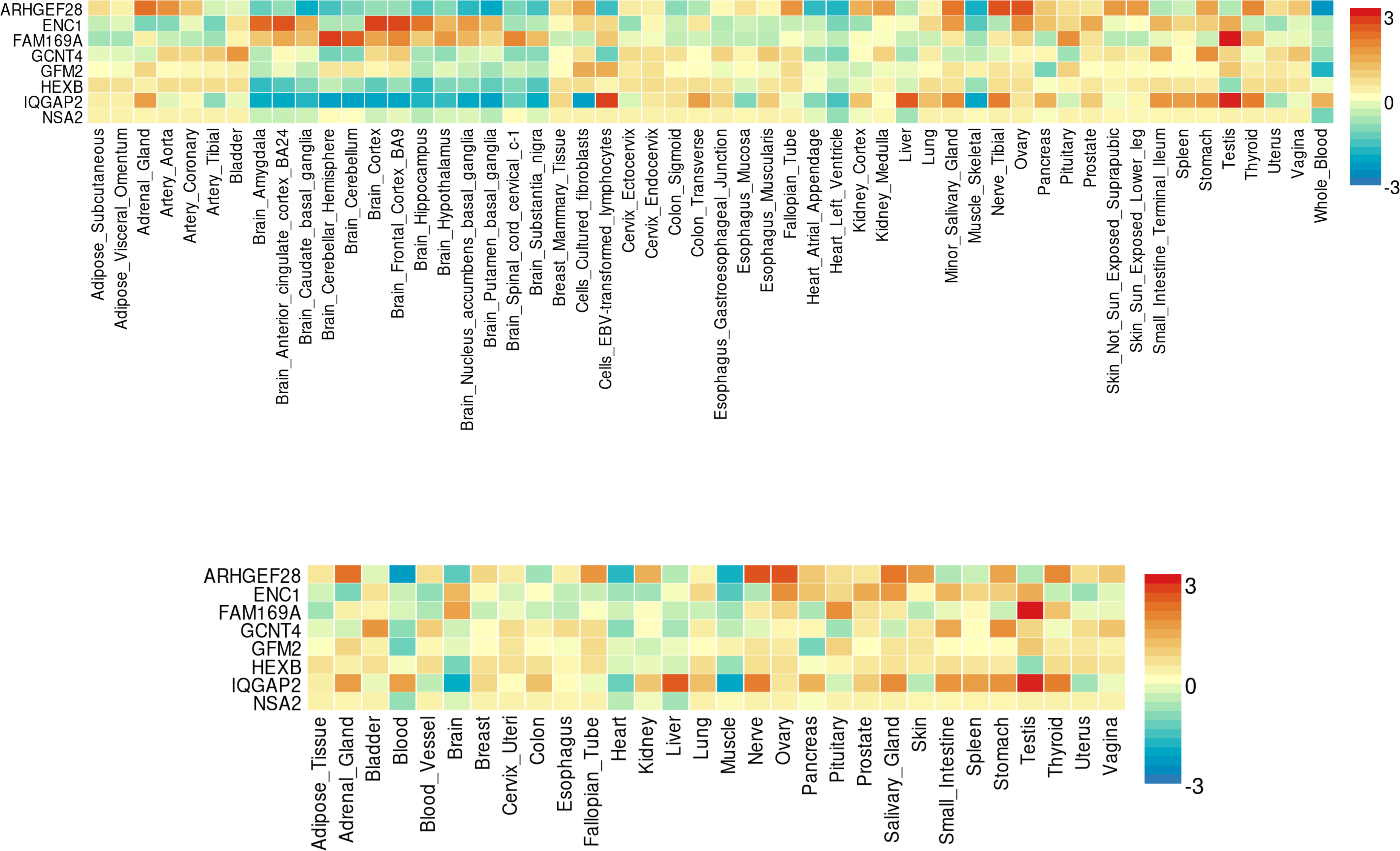
PheWAS results for SNP rs34691025 within primary widespread pain GWAS

**Fig.10.**
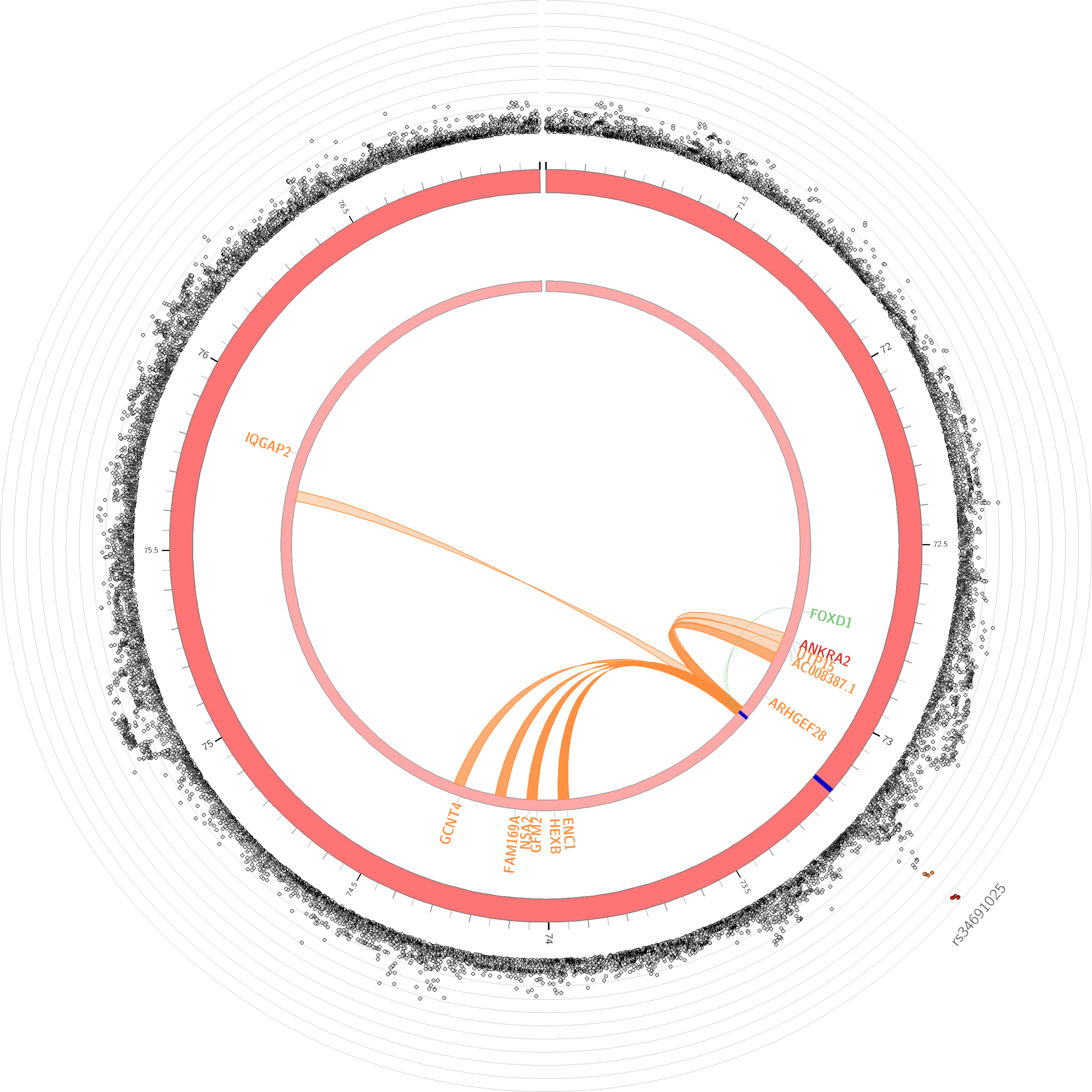
PheWAS results for gene *ARHGEF28* within primary widespread pain GWAS

## Discussion

In this GWAS of widespread pain, conducted using the comprehensive UK Biobank dataset, we identified one novel significant genetic locus and several loci that marginally reached the genome-wide significance level associated with self-reported widespread pain during the past month. In the sex-stratified GWAS, we observed differences in the prevalence of genetic variants of the widespread pain between males and females. Furthermore, our post-GWAS analysis revealed significant genetic correlations between widespread pain and other phenotypes. Additionally, a PheWAS was conducted to further explore the genetic mechanisms underlying widespread pain. In this study, we utilized the generic pain questionnaire from the UK Biobank, a valuable screening instrument, to investigate the potential genetic underpinnings of heterogeneous pain phenotypes, including widespread pain. Employing the UK Biobank offers researchers the advantage of mitigating potential issues related to reduced power due to phenotype heterogeneity. This is achieved by leveraging substantial sample sizes, thereby enhancing the clarity of statistical results amidst potential noise.

In our primary GWAS, we have analyzed and subsequently identified one novel locus that was associated with widespread pain. This significant locus was situated within the *ARHGEF28* gene on chromosome 5q13.2. This particular locus exhibited a significant *p-*value of 1.76 × 10^-8^ for the marker rs34691025. This locus spans an extensive 11 kb within the intronic region of the *ARHGEF28* gene and encompasses as many as 9 genome-wide significant SNPs, as detailed in Supplementary Table 1. The *ARHGEF28* gene (Rho Guanine Nucleotide Exchange Factor 28) is a member of the Rho guanine nucleotide exchange factor family. It plays a pivotal role in regulating Rho GTPases involved in cytoskeletal reorganization and cell migration, may influence pain perception and sensitivity through its effects on neuronal plasticity and inflammatory processes. The cytoskeletal dynamics in neurons, particularly in nociceptive pathways, play a vital role in the development and maintenance of pain states, with alterations in these processes leading to changes in the structural and functional plasticity of pain pathways, thereby impacting pain perception. [21, 22]. Furthermore, *ARHGEF28*’s involvement in immune cell signaling pathways suggests a mechanism by which genetic predispositions could influence the development of chronic pain through immune-mediated inflammation [23, 24]. The link between the immune system and pain has been increasingly recognized, with several studies highlighting how immune responses contribute to the sensitization of the central nervous system, leading to pain conditions [25, 26]. The role of *ARHGEF28* in modulating immune cell functions, including those of mast cells and macrophages, which are key players in inflammation and pain, further supports its potential involvement in pain phenotypes. This dual role in neuronal and immune cell function underscores the complexity of the genetic underpinnings of pain and the need for multidisciplinary approaches to fully understand these mechanisms. The association of *ARHGEF28* with widespread pain underscores the importance of genetic factors in the development of pain disorders and highlights new potential pathways for therapeutic intervention. Further research is needed to explore the functional impact of *ARHGEF28* variations on neuronal and immune cell functions and to understand how these effects contribute to the manifestation of widespread pain. A study identified a heterozygous mutation in amyotrophic lateral sclerosis patients that is predicted to generate a premature truncated *ARHGEF28* gene product, which could provide a possible link between this gene and widespread pain [27]. Moreover, a previous GWAS research has reported a significant association between the *ARHGEF28* gene and hearing problems, suggesting a potential new pathway linking widespread pain and auditory issues [28].

We further identified a cluster of SNPs within the *FAF1* gene on chromosome 1, displaying marginal genome-wide significance, with rs17387024 exhibiting a minimal *p-*value of 7.87 x 10^-8^. The *FAF1* gene, formally known as Fas Associated Factor 1, is a protein-coding gene. The association between the *FAF1* gene and widespread pain may be attributed to its involvement in various biological processes relevant to pain perception and modulation. *FAF1* has been implicated in apoptosis, inflammation, and neuronal signaling, all of which are crucial in the pathogenesis of pain [29]. Studies have shown that *FAF1* is involved in the NF-κB signaling pathway, which plays a significant role in inflammation and pain [30, 31]. Additionally, *FAF1* has been associated with neuronal death and dysfunction, suggesting a potential link to neuropathic pain [32]. The interaction of *FAF1* with other proteins involved in pain pathways, such as *TRPV1*, further supports its role in pain perception [33]. Several previous GWAS have identified the *FAF1* gene as being associated with multisite chronic pain and general pain, providing a possible basis for its link with widespread pain [34-36]. However, further research is still necessary to fully elucidate the exact mechanism by which *FAF1* influences widespread pain.

In the sex-stratified GWAS, we discovered a novel locus unique to the female-specific analysis, located within the *LRMDA* gene on chromosome 10. The top SNP, rs5779595, exhibited a significant *p-*value of 4.04 x 10^-8^. The *LRMDA* gene, also known as leucine-rich melanocyte differentiation-associated protein, has been implicated in some biological processes that could underlie its relationship with pain. Notably, *LRMDA* is involved in melanocyte differentiation and pigmentation, which might suggest modulating nociceptive pathways [37]. Furthermore, several GWAS studies have shown that the *LRMDA* gene is associated with diabetes, a condition that can potentially cause widespread pain and fibromyalgia [38, 39]. This association provides a potential link between the *LRMDA* gene and widespread pain, suggesting that the gene may play a role in the underlying mechanisms of the widespread pain condition. The genetic correlation for widespread pain between males and females was also calculated (*r*_*g*_ = 0.80), indicating a potential disparity in the genetic underpinnings of widespread pain across genders. This divergence is likely attributable to the significant influence of gender differences on the heterogeneity of pain [40]. These findings highlight the necessity of accounting for biological differences between males and females in research on the genetics of pain.

Building on our research findings, there are significant genetic correlations between widespread pain and several other phenotypes, suggesting that shared genetic factors underpinning these conditions. The top three phenotypes most genetically correlated with widespread pain were other joint disorders, not elsewhere classified (*r*_*g*_= 0.94, *p* = 6.20 × 10^-6^), spondylosis (*r*_*g*_ = 0.93, *p* = 7.73 × 10^-6^), and other diseases of the anus and rectum (*r*_*g*_= 0.89, *p* = 1.95 × 10^-6^). Our study provides evidence of the complex interplay of genetic determinants in the manifestation of pain, supporting previous research findings. Specifically, several prior analyses have identified an association between CWP and joint diseases, and some studies suggest that fibromyalgia may share common mechanisms with disorders of the anus and rectum [41, 42]. In the PheWAS, the results reveal that the top SNP (rs34691025) and corresponding gene (*ARHGEF28*) are significantly associated with hearing disorders and cardiovascular diseases. This finding may provide additional evidence that these disorders could be significant contributors to widespread pain, or conversely, that widespread pain may influence the development of these conditions. The shared genetic determinants, correlations with other pain phenotypes, and associations with various medical conditions and medications highlight the necessity of adopting a holistic approach to comprehensively understanding and addressing widespread pain.

In our research, we employed a novel operational definition to delineate cases of widespread pain, characterizing them as individuals experiencing pain throughout their body during the last month. This approach stands in contrast to previous GWAS that defined cases based on the presence of pain all over the body lasting for more than three months [9]. Notably, the CWP cases in these studies numbered 6,914, a criterion that differs from our own. The adoption of this alternative definition was driven by the consideration that a one-month duration might capture a broader spectrum of pain phenotypes, potentially including both acute and chronic manifestations. By focusing on a one-month timeframe, we intend to provide a distinct and meaningful delineation of widespread pain cases, offering a new perspective on the genetic underpinnings of pain phenotypes. Our choice of the definition is motivated by the goal of enhancing the understanding of genetic factors associated with widespread pain.

Our research provides initial insights into the genetic underpinnings of widespread pain. However, it encounters certain limitations, particularly in the validation phase of our results. During the replication phase, we utilized three public datasets of CWP and fibromyalgia, yet we did not achieve significant replication results for our study. This outcome could potentially be attributed to differences in the definitions of widespread pain, CWP, and fibromyalgia. Additionally, the limited number of participants in the replication cohorts might have contributed to this challenge. In the female-specific GWAS, rs5779595 becomes the top and only SNP identified within the locus, pointing to the potential spuriousness of this association and underscoring the need for further exploration and study. Furthermore, it is essential to discuss the methodological approach adopted in our study. We characterized widespread pain cases and controls based on the responses provided by UK Biobank participants to a specific question regarding pain experienced over the last month. However, a critical observation is that this question did not delve into nuanced details such as the severity or frequency of the pain. Consequently, the phenotype definition derived from these responses should be perceived and interpreted as being broadly defined. This broad definition may have implications for the specificity and sensitivity of our findings, as it potentially includes a heterogeneous group of individuals with varying pain experiences. Future research may need to incorporate more detailed questionnaires and larger sample sizes to capture the complexity of widespread pain and provide a more comprehensive understanding of its genetic basis. Additionally, exploring alternative phenotype definitions that account for pain severity and frequency could enhance the precision of genetic associations and contribute to a deeper understanding of the mechanisms underlying widespread pain.

## Conclusion

In conclusion, our primary GWAS of widespread pain, utilizing the extensive UK Biobank dataset, successfully identified one novel genetic locus and several loci that reached genome-wide significance. The sex-stratified GWAS outputs revealed gender-specific variants associated with of widespread pain between males and females. Furthermore, our post-GWAS analysis demonstrated substantial genetic correlations and shared mechanisms between widespread pain and other phenotypes. This study advances our understanding of the genetic factors contributing to widespread pain.

## Declarations

## Data availability

The summary statistics of the UK Biobank results on widespread pain can be accessed upon publication. Any other data relevant to the study that are not included in the article or its supplementary materials are available from the authors upon reasonable request.

## Data Availability

All data produced in the present study are available upon reasonable request to the authors
All data produced in the present work are contained in the manuscript

## Acknowledgements

The authors extend their sincere gratitude to the participants of the UK Biobank, FinnGen, FibroGene, and TwinsUK cohorts for their invaluable contributions of genetic and phenotypic data. This study will adhere to all ethical guidelines and data protection protocols of the UK Biobank. This research has been conducted using the UK Biobank Resource under Application Number 89386.

## Authors’ contributions

QP drafted the paper and performed the UK Biobank GWAS analysis. TC and YT contributed to data formatting. MH and TD provided comments to the paper. RC, MKN and FMKW conducted the replication study and provided comments to the paper. WM organized the project and provided comments.

## Corresponding authors

Correspondence to Weihua Meng

## Consent for publication

All authors have consent for publication.

## Ethics approval and consent to participate

This study was approved by the Ethics Committee of the University of Nottingham, Ningbo, China. All authors have consent for participation. The TwinsUK Study was approved by London-Westminster Research Ethics Committee (REC referenceEC04/015), and Guy’s and St Thomas’ NHS Foundation Trust Research and Development (R&D). The TwinsUK Biobank was approved by the HRA - Liverpool East Research Ethics Committee (REC reference 19/NW/0187), IRAS ID 258513. All participants provide written, informed consent.

