## Supplementary figures and images for "Genome-Wide Association Study Identifies Novel Genetic Variants Associated with Widespread Pain in the UK Biobank (N=172,230)"

### Supplementary Figure 1

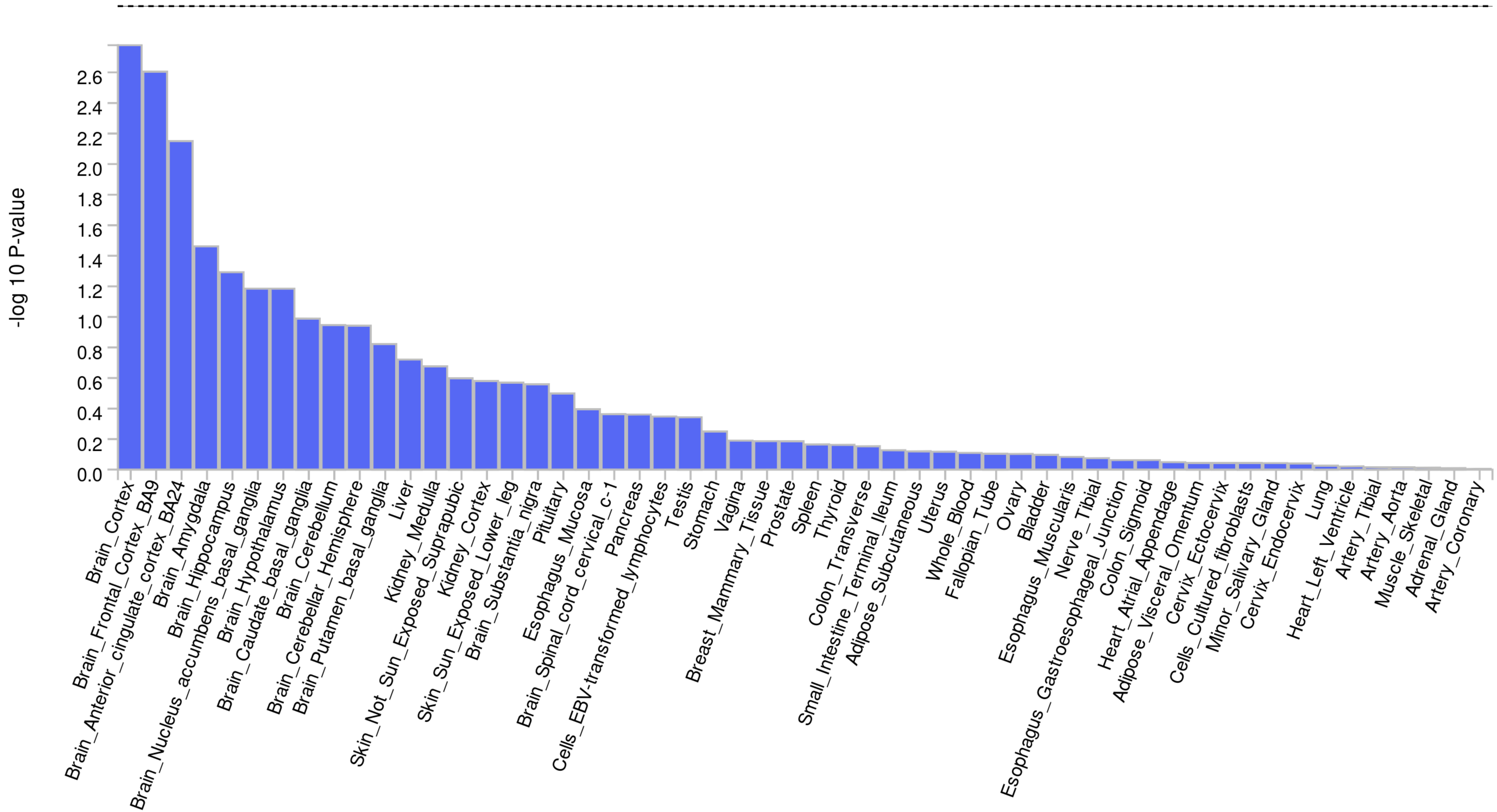

### Supplementary Figure 2

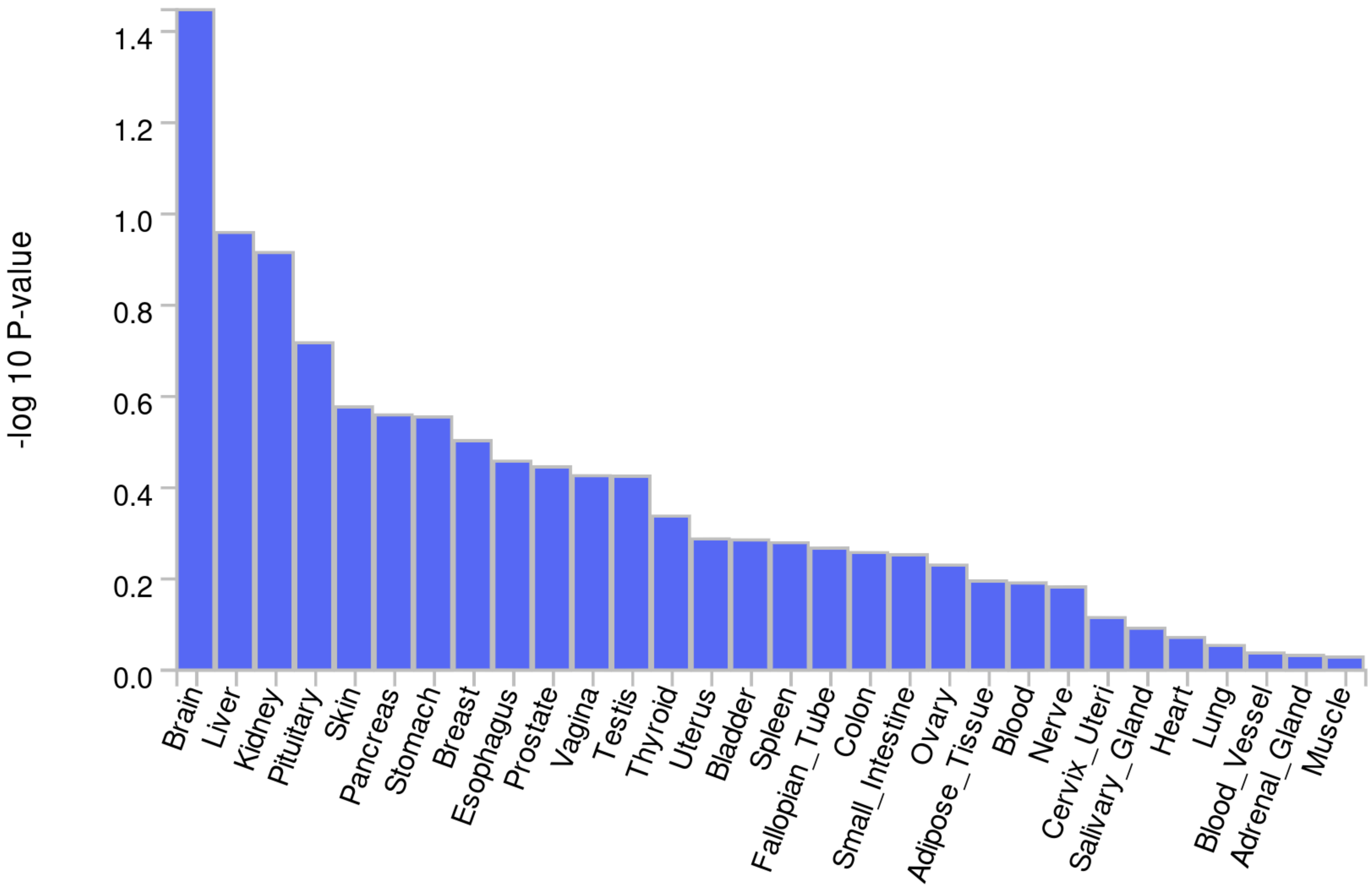

### Supplementary Figure 3

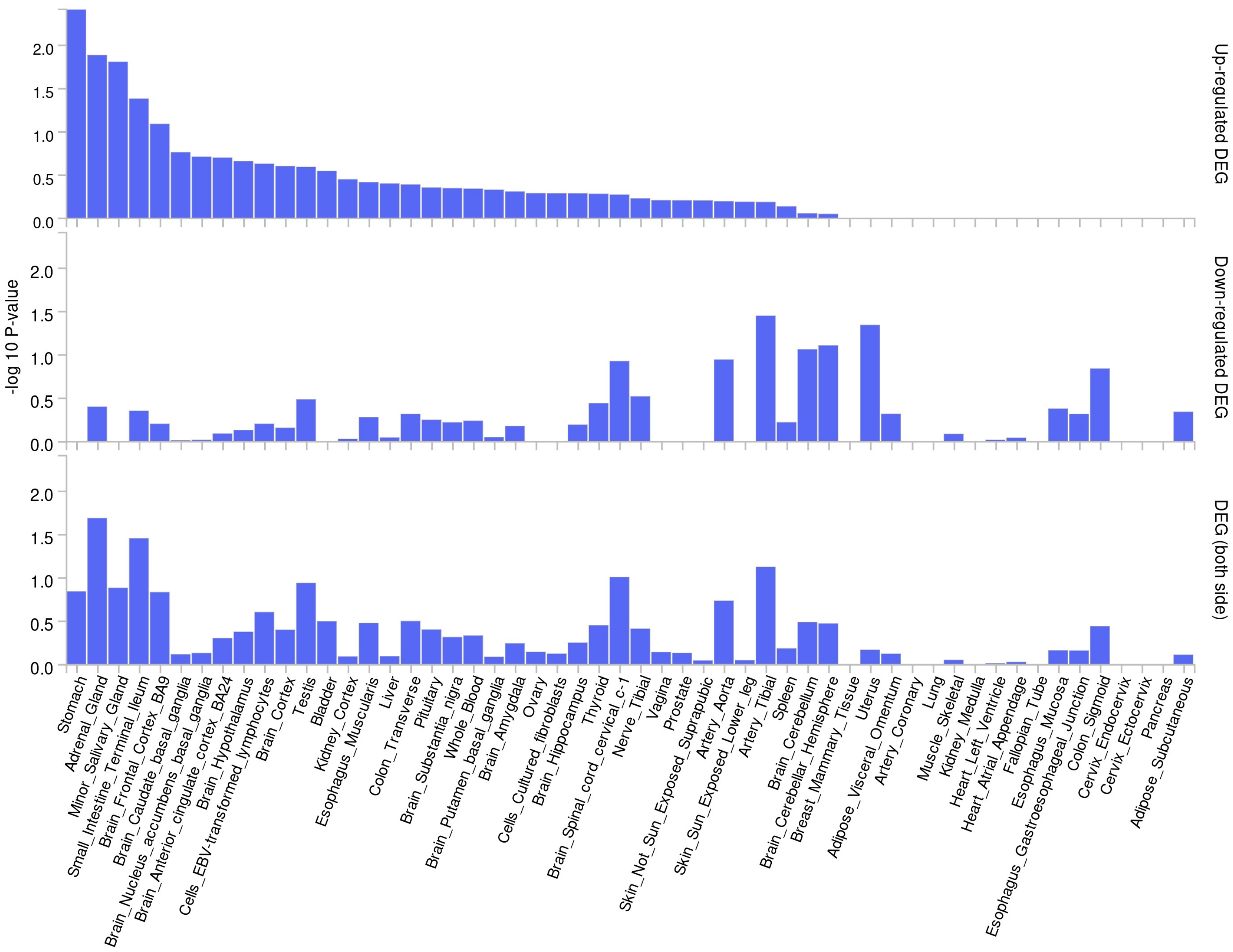

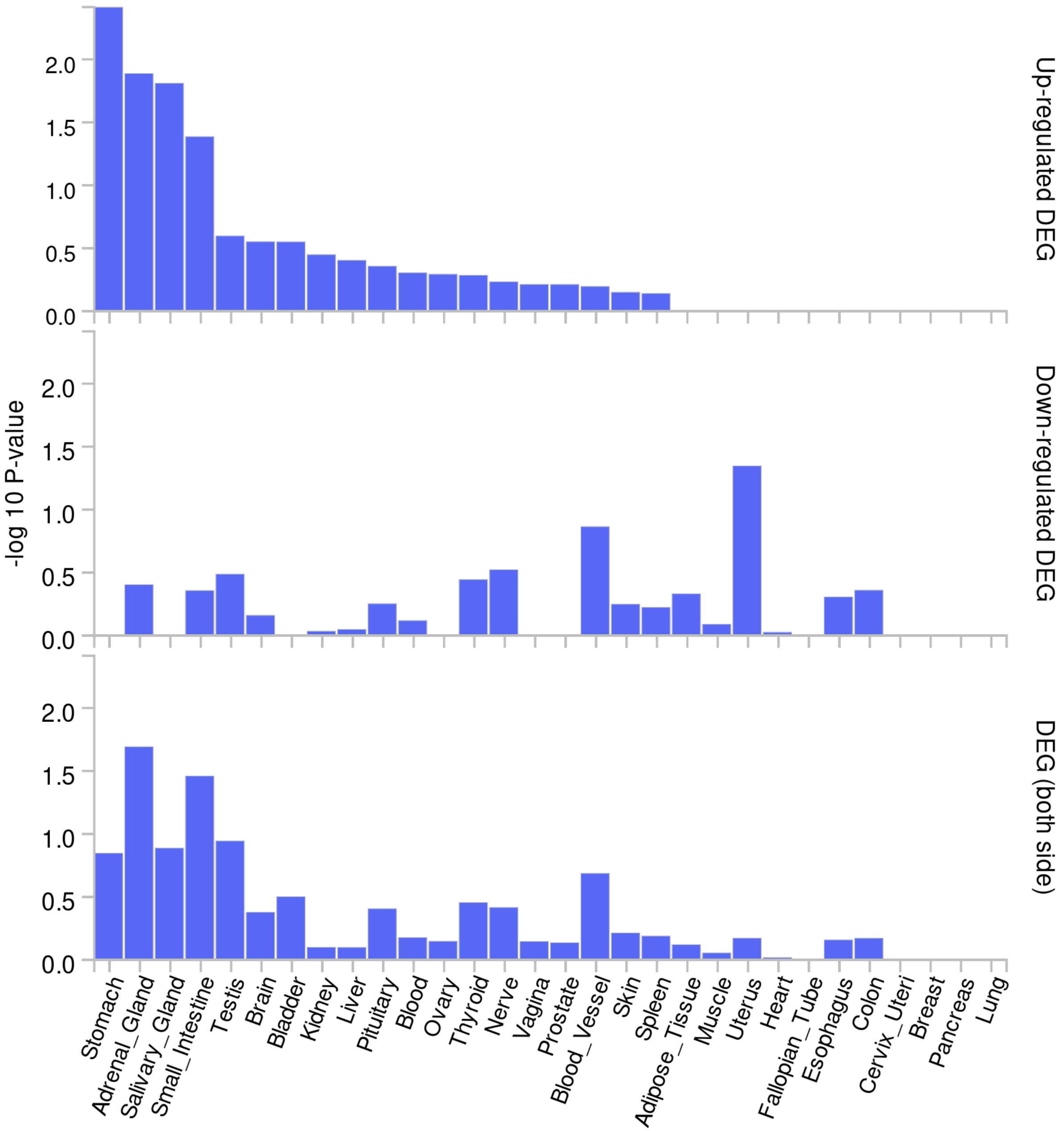
